# Contribution of genetics and lifestyle to the risk of major cardiovascular and thromboembolic complications following COVID-19

**DOI:** 10.1101/2022.10.26.22281547

**Authors:** Junqing Xie, Yuliang Feng, Danielle Newby, Bang Zheng, Qi Feng, Albert Prats-Uribe, Chunxiao Li, Nicholas J Wareham, R Paredes, Daniel Prieto-Alhambra

## Abstract

Clinical determinants for cardiovascular and thromboembolic (CVE) complications of COVID-19 are well-understood, but the roles of genetics and lifestyle remain unknown. We performed a prospective cohort study using UK Biobank, including 25,335 participants with confirmed SARS-CoV-2 infection between March 1, 2020, and September 3, 2021. Outcomes were hospital-diagnosed atrial fibrillation (AF), coronary artery disease (CAD), ischemic stroke (ISS), and venous thromboembolism (VTE) within 90 days post-infection. Heritable risk was represented by validated polygenic risk scores (PRSs). Lifestyle was defined by a composite of nine variables. We estimated adjusted hazard ratios (aHR) and confidence intervals (CI) using Cox proportional hazards models. In the COVID-19 acute phase, PRSs linearly predicted a higher risk of AF (aHR 1.52 per standard deviation increase, 95% CI 1.39 to 1.67), CAD (1.59, 1.40 to 1.81), and VTE (1.30, 1.11 to 1.53), but not ISS (0.92, 0.64 to 1.33). A healthy lifestyle was associated with a substantially lower risk of post-COVID-19 AF (0.70, 0.53 to 0.92), CAD (0.64, 0.44 to 0.91), and ISS (0.28, 0.12 to0.64), but not VTE (0.82, 0.48 to 1.39), compared with an unhealthy lifestyle. No evidence for interactions between genetics and lifestyle was found. Our results demonstrated that population genetics and lifestyle considerably influence cardiovascular complications following COVID-19, with implications for future personalised thromboprophylaxis and healthy lifestyle campaigns to offset the elevated cardiovascular disease burden imposed by the ongoing pandemic.

## Introduction

Cardiovascular disease is the leading cause of death globally.^1^ Recently, cardiovascular mortality and morbidity have risen further due to the direct and indirect consequences of the COVID-19 pandemic.^2,3^ It is expected that the repercussions and long-term sequelae of COVID-19 could further increase the cardiovascular burden to an unprecedented level.^4^

At the individual level, preventing life-threatening cardiovascular and thromboembolic events (CVE) is crucial during the treatment of patients with COVID-19. However, a challenge remains to accurately identify individuals at sufficient risk to warrant intensive surveillance or targeted pharmacological prophylaxis. For instance, although prophylactic anticoagulation has been recommended for hospitalized patients with COVID-19^5^, evidence for its use is vastly conflicting for more critical ICU patients and milder ambulatory patients with COVID-19^6–8^.

General risk factors, including age, sex, and obesity, are recognized predictors of COVID-19 severity, including hospital admission and mechanical ventilation. However, although they are helpful in informing clinical practice, they are not specific to CVE risk, and commonly correlate with an elevated risk of bleeding. In contrast, polygenic risk scores (PRSs), a sum of genetic risk for a particular trait, have been proposed as a promising tool for precision medicine and early cardiovascular risk stratification.^9–11^ It is not yet known whether this previously well-captured genetic susceptibility to typical CVE will also predict the occurrence of COVID-19-related CVE during acute and post-acute illness phases.

In addition, effective public health interventions are urgently needed to reduce the population’s cardiovascular burden, particularly in light of surging COVID-19 infections after the removal of most early restrictions (e.g., lockdown and social distance). The US Preventive Service Task Force updated its recommendations in 2022, promoting healthy behaviour counselling for all adults as a national strategy for primary cardiovascular prevention.^12^ However, all current clinical and public health guidelines^13,14^ lack insights into the potential role of healthy lifestyle modifications in alleviating COVID-19 cardiovascular complications, likely due to a paucity of evidence.

We aimed to assess the association between validated PRSs, lifestyle risk factors, and their interactions and the risk of major CVE within 90 days after COVID-19 diagnosis.

## Results

### Population characteristics

Table 1 shows the baseline characteristics of the whole UK Biobank eligible source population with available PRS (n=408,395) and of participants with COVID-19 who were eligible for analyses (n=25,335), overall and stratified by their genetic risk for AF. The average (SD) age of the COVID-19 cohort was 65.99 (8.53) years, of whom most were women (52.7%) and of White ethnicity (84.6%). The prevalence of the nine prespecified unhealthy lifestyle factors ranged from 10.8% for smoking status to 48.5% for low oily fish intake. In total, 8.6% of infected individuals lived an unhealthy lifestyle based on our composite measure (>=5 unhealthy lifestyle factors), a proportion generally consistent with that of the entire UK Biobank source population. All study covariates, except ethnicity, were distributed similarly across different genetic strata, illustrating independence between the selected PRS and lifestyle factors. The baseline characteristics stratified by other PRSs for CAD, ISS, and VTE were similar to those when stratified by the AF genetic score (see Supplementary Table 1–3).

**Table 1:** Characteristics of the whole UK Biobank population and of participants with COVID-19, stratified by genetic risk of atrial fibrillation.^1^.

| Participant characteristics | All eligible UK Biobank participants<sup>2</sup> | Participants with COVID-19 |  |  |  |
| --- | --- | --- | --- | --- | --- |
|  |  | All | Low genetic risk | Intermediate genetic risk | High genetic risk |
| Number | 408395 | 25335 | 5067 | 15201 | 5067 |
| Mean age, year (SD) | 68.10 (8.10) | 65.99 (8.53) | 65.94 (8.52) | 66.06 (8.53) | 65.82 (8.56) |
| sex, No. (%) |  |  |  |  |  |
| Female | 225451 (55.2) | 13339 (52.7) | 2647 (52.2) | 7984 (52.5) | 2708 (53.4) |
| Male | 182944 (44.8) | 11996 (47.3) | 2420 (47.8) | 7217 (47.5) | 2359 (46.6) |
| Ethnicity, No. (%) |  |  |  |  |  |
| White | 382169 (93.6) | 3893 (15.4) | 939 (18.5) | 2189 (14.4) | 765 (15.1) |
| Other ethnic groups | 26226 ( 6.4) | 21442 (84.6) | 4128 (81.5) | 13012 (85.6) | 4302 (84.9) |
| Obesity, No. (%) |  |  |  |  |  |
| BMI < 30 | 311205 (76.2) | 17984 (71.0) | 3643 (71.9) | 10834 (71.3) | 3507 (69.2) |
| BMI ≥ 30 | 97190 (23.8) | 7351 (29.0) | 1424 (28.1) | 4367 (28.7) | 1560 (30.8) |
| Education categories<sup>3</sup>, No. (%) |  |  |  |  |  |
| I | 73987 (18.1) | 4863 (19.2) | 940 (18.6) | 2920 (19.2) | 1003 (19.8) |
| II | 110969 (27.2) | 7999 (31.6) | 1592 (31.4) | 4822 (31.7) | 1585 (31.3) |
| III | 45409 (11.1) | 2613 (10.3) | 492 ( 9.7) | 1570 (10.3) | 551 (10.9) |
| IV | 20765 ( 5.1) | 1194 ( 4.7) | 251 ( 5.0) | 701 ( 4.6) | 242 ( 4.8) |
| V | 157265 (38.5) | 8666 (34.2) | 1792 (35.4) | 5188 (34.1) | 1686 (33.3) |
| Index of multiple deprivation, mean (SD) | 17.51 (13.87) | 20.62 (14.85) | 20.51 (14.78) | 20.60 (14.89) | 20.77 (14.80) |
| **Individual lifestyle factors, No. (%)** |  |  |  |  |  |
| Smoking status |  |  |  |  |  |
| Never or past | 368471 (90.2) | 22590 (89.2) | 4529 (89.4) | 13556 (89.2) | 4505 (88.9) |
| Current | 39924 ( 9.8) | 2745 (10.8) | 538 (10.6) | 1645 (10.8) | 562 (11.1) |
| Alcohol intake |  |  |  |  |  |
| ≤ 4 times week | 325341 (79.7) | 21062 (83.1) | 4228 (83.4) | 12621 (83.0) | 4213 (83.1) |
| Daily or almost daily | 83054 (20.3) | 4273 (16.9) | 839 (16.6) | 2580 (17.0) | 854 (16.9) |
| Physical activity |  |  |  |  |  |
| ≥150 min per week of MIPA or ≥75 min per week of VIPA | 345208 (84.5) | 21194 (83.7) | 4229 (83.5) | 12753 (83.9) | 4212 (83.1) |
| <150 min per week of MIPA & <75 min per week of VIPA | 63187 (15.5) | 4141 (16.3) | 838 (16.5) | 2448 (16.1) | 855 (16.9) |
| Television viewing time |  |  |  |  |  |
| < 4 h/day | 293476 (71.9) | 17880 (70.6) | 3630 (71.6) | 10708 (70.4) | 3542 (69.9) |
| ≥ 4 h/day | 114919 (28.1) | 7455 (29.4) | 1437 (28.4) | 4493 (29.6) | 1525 (30.1) |
| Sleep duration |  |  |  |  |  |
| ≥7 & ≤9h/day | 301723 (73.9) | 18121 (71.5) | 3626 (71.6) | 10879 (71.6) | 3616 (71.4) |
| <7 or >9h/day | 106672 (26.1) | 7214 (28.5) | 1441 (28.4) | 4322 (28.4) | 1451 (28.6) |
| Fruit and vegetable intake |  |  |  |  |  |
| ≥ 400 g/day | 327452 (80.2) | 19456 (76.8) | 3889 (76.8) | 11649 (76.6) | 3918 (77.3) |
| <400 g/ day | 80943 (19.8) | 5879 (23.2) | 1178 (23.2) | 3552 (23.4) | 1149 (22.7) |
| Oily fish intake |  |  |  |  |  |
| ≥1 portion/week | 230406 (56.4) | 13047 (51.5) | 2633 (52.0) | 7802 (51.3) | 2612 (51.5) |
| <1 portion/week | 177989 (43.6) | 12288 (48.5) | 2434 (48.0) | 7399 (48.7) | 2455 (48.5) |
| Red meat intake |  |  |  |  |  |
| ≤3 portion/week | 188229 (46.1) | 13118 (51.8) | 2576 (50.8) | 7834 (51.5) | 2708 (53.4) |
| >3 portion/week | 220166 (53.9) | 12217 (48.2) | 2491 (49.2) | 7367 (48.5) | 2359 (46.6) |
| Processed meat intake |  |  |  |  |  |
| ≤1 portion/week | 282323 (69.1) | 16716 (66.0) | 3329 (65.7) | 10037 (66.0) | 3350 (66.1) |
| >1 portion/week | 126072 (30.9) | 8619 (34.0) | 1738 (34.3) | 5164 (34.0) | 1717 (33.9) |
| Combined lifestyle, No. (%) |  |  |  |  |  |
| Healthy (0-4 unhealthy factors) | 377279 (92.4) | 23151 (91.4) | 4624 (91.3) | 13884 (91.3) | 4643 (91.6) |
| Unhealthy (5-9 unhealthy factors) | 31116 ( 7.6) | 2184 ( 8.6) | 443 ( 8.7) | 1317 ( 8.7) | 424 ( 8.4) |

CAD: Coronary artery disease, BMI: Body mass index.
<sup>1</sup> The baseline characteristics stratified by genetic risk of coronary artery disease, ischaemic stroke, and venous thromboembolism were similar to that of atrial fibrillation. <sup>2</sup> All eligible participants in UK Biobank included those who survived when this study began (March 1, 2020). <sup>3</sup> I include self-reported “None of the above” and “Prefer not to answer”, II includes “CSEs or equivalent” and “O levels/GCSEs or equivalent”, III includes “A levels/AS levels or equivalent”, IV includes “Other professional qualifications e.g.: nursing, teaching”, and V includes “NVQ or HND or HNC or equivalent” and “College or University degree.”

After infection, 422 AF, 244 CAD, 29 ISS, and 135 VTE events occurred during the 90-day follow-up period, accounting for 1.67%, 0.96%, 0.12%, and 0.53%, of the COVID-19 cohort, respectively. The incidence rate was 6.12 per 1,000 person-years for ISS, 28.0 for VTE, 48.5 for CAD, and 86.9 for AF.

### Genetic risk and post-COVID-19-CVE

All four PRSs followed a normal distribution (see Supplementary Figure 1). A higher PRS for CAD, AF, or VTE was associated with an increased risk of post-COVID-19 CVE, with an HR per SD increase in the PRS of 1.52 (95% CI 1.39 to 1.67) for AF, 1.59 (1.40 to 1.81) for CAD, and 1.30 (1.11 to 1.53) for VTE (Table 2). These associations were significantly linear, with all P-values for the linear term < 0.001 and for the non-linear term >0.05 (see Supplementary Figure 1). There was no association between ISS-PRS and post-COVID-19 ISS (HR: 0.92 [0.64 to 1.33]).

**Table 2:** Associations between polygenic risk scores (PRSs) and 90-day risk of cardiovascular and thromboembolic events after COVID-19.

| Outcomes | No. of events | Incidence (per 1,000 pys) | Adjusted hazard ratio* (95% CI) |  |  |  | *P*-trend |
| --- | --- | --- | --- | --- | --- | --- | --- |
|  |  |  | Continuous PRS (per SD increase) | Low genetic risk | Intermediate genetic risk | High genetic risk |  |
| Atrial fibrillation | 422 | 86.9 | 1.52 (1.39 to 1.67) | 1 (ref) | 1.57 (1.15 to 2.15) | 3.00 (2.16 to 4.15) | <0.01 |
| Coronary artery disease | 235 | 48.5 | 1.59 (1.40 to 1.81) | 1 (ref) | 1.53 (1.00 to 2.34) | 3.45 (2.22 to 5.34) | <0.01 |
| Ischaemic stroke | 29 | 6.12 | 0.92 (0.64 to 1.33) | 1 (ref) | 0.96 (0.38 to 2.44) | 0.79 (0.24 to 2.60) | 0.85 |
| Venous thromboembolism | 135 | 28.0 | 1.30 (1.11 to 1.53) | 1 (ref) | 1.22 (0.75 to 2.00) | 2.00 (1.15 to 3.38) | <0.01 |

Pys: person-years, Ref: reference, SD: standard deviation

Figure 1 depicts Kaplan-Meier cumulative incidence curves and shows that CVE increased dramatically within the first 15 to 30 days after infection. Early separation of risk was consistently observed across the three genetic risk categories and continued to diverge over time. Compared to participants with low genetic risk, those with higher genetic risk had a significantly increased incidence of CVE post-COVID-19. The HR of high vs low genetic risk was 3.00 (2.16 to 4.15) for AF, 3.45 (2.22 to 5.34) for CAD, and 2.00 (1.15 to 3.38) for VTE.

**Figure 1:**
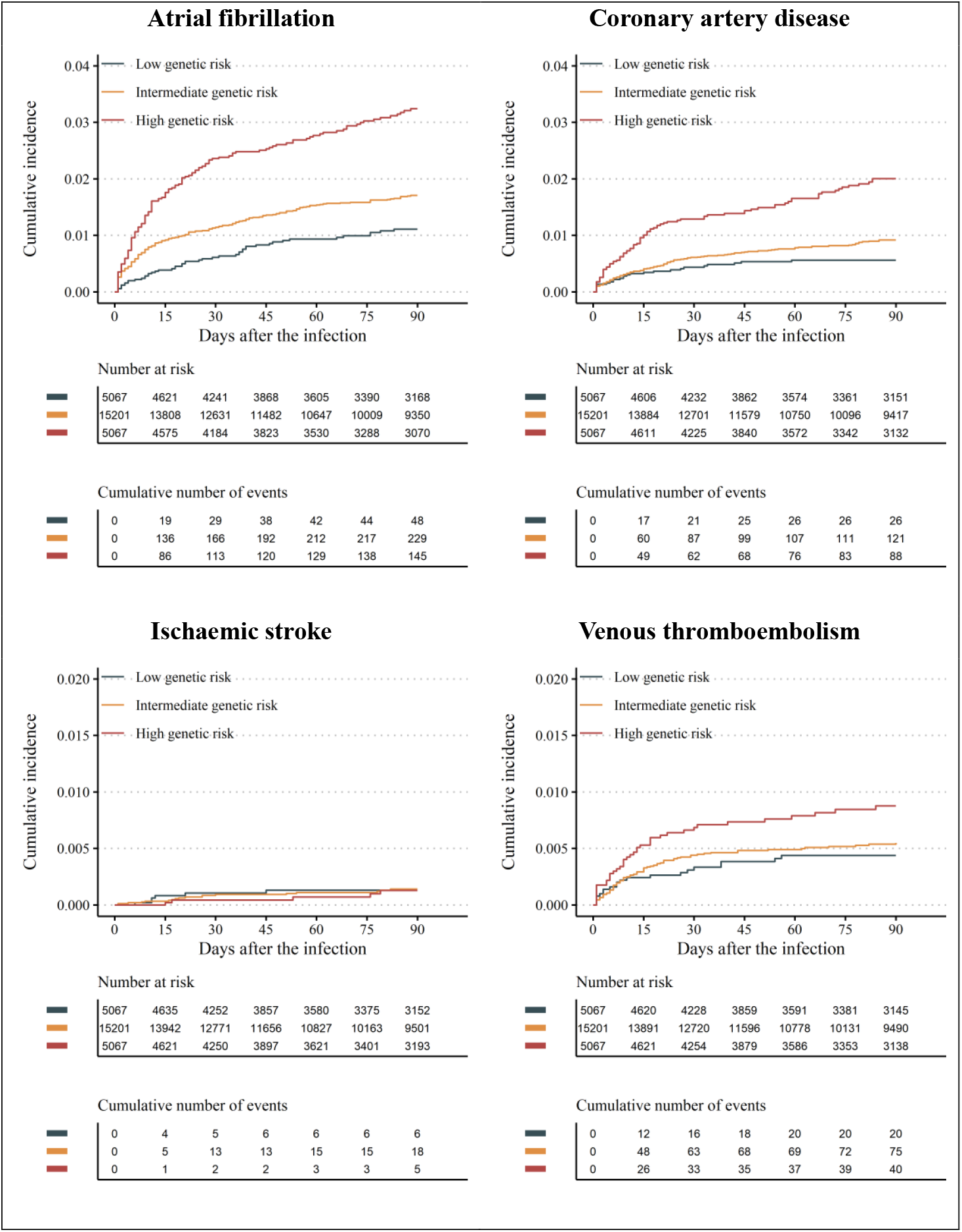
Cumulative incidence of COVID-19-related cardiovascular events by the three genetic risk categories. The upper limit of the y-axis was 0.04 for atrial fibrillation and coronary artery disease and was 0.02 for ischaemic stroke and venous thromboembolism.

The overall estimate of the association between each PRS and AF, CAD, and VTE was similar across a range of prespecified subgroups, despite the substantially varying baseline risks (Figure 2). For example, the incidence rate of CAD among participants with COVID-19 aged 65 years or older was 78.03 per 1,000 person-years, almost 5-fold higher than those younger than 65 years (incidence rate: 19.64), but the HR estimate was similar (1.57 for those aged over 65 years vs 1.67 for those aged under 65 years, P-interaction = 0.61). Recent users of antithrombotics had substantially higher background risks of CAD and AF, with incidence rate of 264.99 per 1,000 person-years for users and 12.07 for non-users for CAD and incidence rates of 338.42 for users and 44.78 for non-users for AF. The association between PRS and post-COVID-19 CVE was attenuated, but remained significant (HR: 1.20 for those antithrombotic users vs 1.99 for those non-antithrombotic users for CAD, P-interaction < 0.01; 1.34 vs 1.63 for AF, P-interaction= 0.05). The genetic risk appeared to persist among fully vaccinated individuals (HR: 1.62 for breakthrough infection vs 1.50 for non-breakthrough infection for AF, P-interaction = 0.54; 1.23 vs 1.63 for CAD, P-interaction = 0.18; 1.51 vs 1.29 for VTE, P-interaction = 0.62).

**Figure 2:**
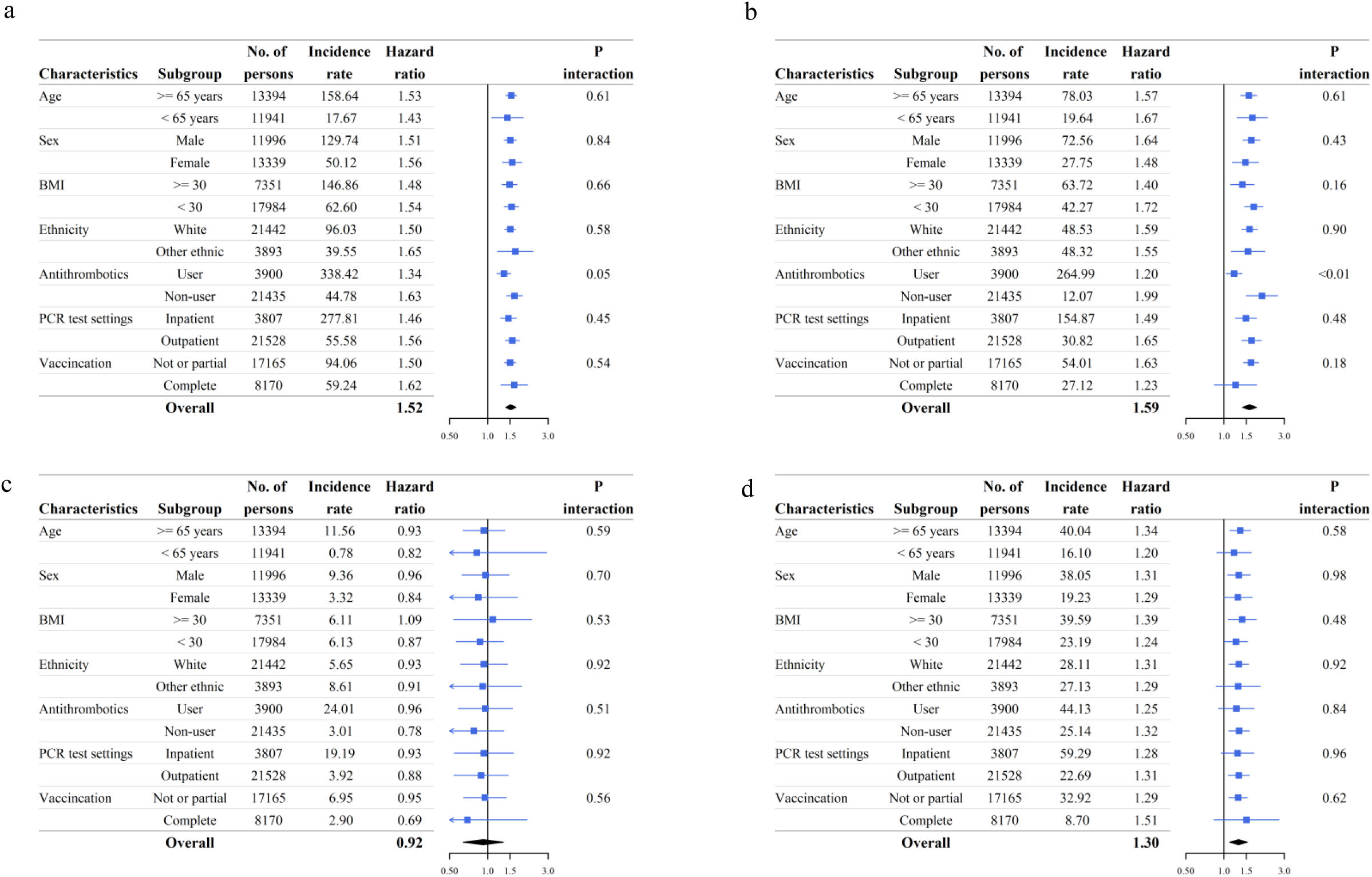
Genetic risk of COVID-19-related cardiovascular events across clinically relevant subgroups. a: atrial fibrillation, b: coronary artery disease, c: ischaemic stroke, d: venous thromboembolism.

### Healthy lifestyle and COVID-19-related CVE

Compared to those with an unhealthy lifestyle, people adhering to healthier habits had a significantly reduced risk of AF (HR 0.70 [95% CI 0.53 to 0.92]), CAD (0.64 [0.44 to 0.91]), and ISS (0.28 [0.12 to 0.64]) within 90 days following COVID-19 infection. No association was found for post-COVID-19 VTE [0.82 (0.48 to 1.39)]. With 8.6% of infected individuals having an unhealthy lifestyle, the prevented fraction of CVE outcomes for the COVID-19 population changing to a healthy lifestyle was 2.46% for AF, 2.95% for CAD, and 5.86% for ISS (Table 3). There was no significant interaction between genetic and lifestyle factors for any CVE outcomes (P-interaction > 0.05). However, there was an additive effect for the risk of AF and CAD, with an HR for high genetic risk and unhealthy lifestyle vs low genetic risk and healthy lifestyle of 5.09 [3.07 to 8.41] for AF and 5.00 [2.52 to 9.93] for CAD (Figure 3).

**Table 3:** Hazard ratios for COVID-19-related cardiovascular and thromboembolic events and prevented fraction of the population associated with a combined healthy lifestyle.^1^.

| Outcomes | Hazard ratio (healthy lifestyle vs unhealthy lifestyle) |  |  |  |  | Prevented fraction of the population (%)<sup>2</sup> |
| --- | --- | --- | --- | --- | --- | --- |
|  | All participants with COVID-19 | Low genetic risk | Intermediate genetic risk | High genetic risk | P-interaction |  |
| Atrial fibrillation | 0.70 (0.53 to 0.92) | 1.28 (0.49 to 3.30) | 0.73 (0.49 to 1.07) | 0.52 (0.33 to 0.81) | 0.10 | 2.46 |
| Coronary artery disease | 0.64 (0.44 to 0.91) | 1.03 (0.30 to 3.56) | 0.57 (0.35 to 0.92) | 0.63 (0.34 to 1.15) | 0.96 | 2.95 |
| Ischaemic stroke | 0.28 (0.12 to 0.64) | 0.25 (0.02 to 3.05) | 0.19 (0.07 to 0.52) | 0.28 (0.01 to 13.4) | 0.99 | 5.86 |
| Venous thromboembolism | 0.82 (0.48 to 1.39) | 0.43 (0.14 to 1.33) | 0.77 (0.38 to 1.57) | 1.59 (0.48 to 5.21) | 0.07 | NA |

<sup>1</sup> 8.1% of people in the COVID-19 cohort recorded an unhealthy lifestyle. <sup>2</sup>Prevented fraction for the population was calculated according to the formula: PFP = $P_e (1 - HR)$ , where $P_e$ was the prevalence of an unhealthy lifestyle<sup>45</sup>.

**Figure 3:**
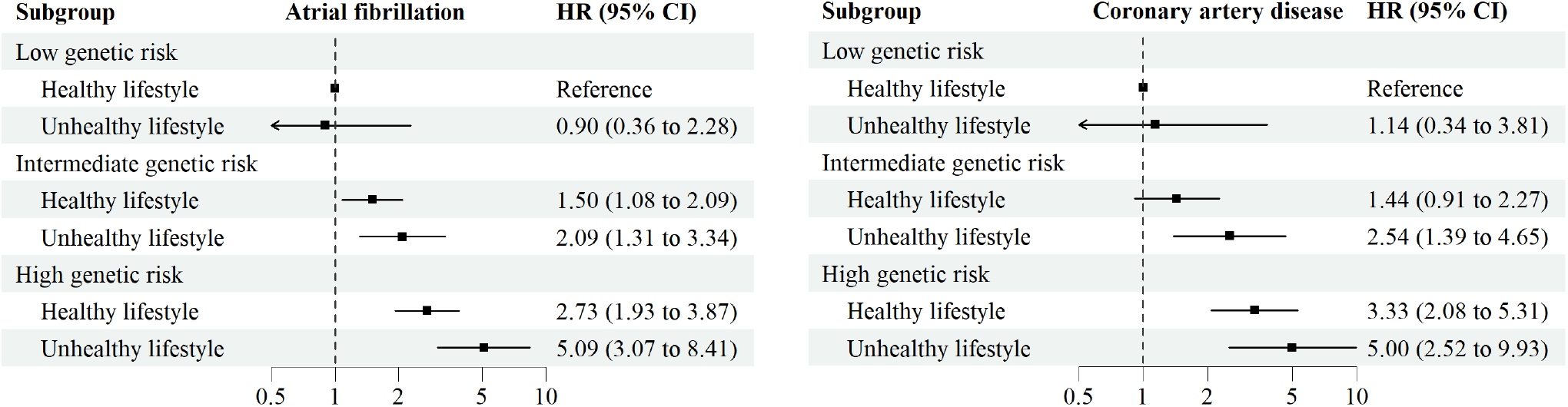
Combined effect of genetic and lifestyle factors on COVID-19-related atrial fibrillation and coronary arterial disease. HR: hazard ratio. We only examined joint effects for cardiovascular events that had significant associations with both genetic and lifestyle factors in the primary analysis.

### Sensitivity analyses

The results of the sensitivity analysis were generally consistent with the primary analysis. Noticeably, the magnitude of PRS association was reduced for the sensitivity analysis of incident events (HR: AF 1.33 [1.11 to 1.59]; CAD 1.19 [0.87 to 1.63]) and hospital admission-related events (AF 1.31 [0.89 to 1.92]; CAD 1.21 [0.71 to 2.08]), compared to those estimates regarding any events diagnosed after infection. However, the genetic risk was even more pronounced when restricting to admission-related VTE outcome [HR 1.52 (1.14 to 2.02)].

As expected, the positive control experiment shown that PRSs were prospectively associated with the corresponding CVE event among participants without COVID-19. And the negative control experiment found that the post-COVID-19 risk of diabetes was not associated with an increase of most of PRSs, except for AF-PRS. (Supplementary Table 4)

## Discussion

### Key findings

This is the first prospective cohort study of the genetic- and lifestyle-related determinants of post-COVID-19 CVE complications. We found that a higher genetic risk based on validated PRSs for CVE events was linearly associated with an increased incidence of post-COVID-19 CVE. Participants with the top 20% of PRSs had a 3-fold, 3.5-fold, and 2-fold excess risk of AF, CAD, and VTE, respectively, compared with those with the lowest 20% of PRSs. No significant association was observed for post-COVID-19 ISS. The identified genetic predisposition was persistent in subgroups of participants who were already at very high risk of CVE and were receiving antithrombotic therapy before infection and in those with a breakthrough infection after full vaccination (2 doses).

We demonstrated that a healthy lifestyle was associated with substantially lower risk of arterial events (−30% AF, −36% CAD, and −72% ISS) among participants with COVID-19. A sum of over 10% of CVE (2.46% AF, 2.95% CAD, 5.86% VTE) could have been prevented if all individuals with an unhealthy lifestyle had adopted a healthier one. We did not observe a significantly different association between healthy lifestyle and CVE risk across different genetic risk strata. Instead, high genetic risk and unhealthy behaviour had an additive effect on the increased risk of post-COVID-19 AF and CAD complications.

### Findings in context

Our study is the first to investigate genetic determinants for COVID-19-related CVE. It differs from previous studies of PRS for adult-onset cardiovascular diseases^15–18^ as we targeted people with COVID-19 rather than the general healthy population and we evaluated the risk of acute CVE triggered by SARS-CoV-2 rather than the chronic (≥10 years) or lifetime disease risk.

The strongest associations (HR per one SD increase) found in previous studies were 2.33 for AF^16^, 1.86 for CAD^16^, 1.26 for ISS^19^, and 1.27 for VTE^18^, compared with 1.52 for AF, 1.59 for CAD, 0.96 for ISS, and 1.30 for VTE found here. Our observed associations demonstrate that population genetic variations are an important contributor to developing CVE following COVID-19 and highlight underlying genetic interconnections between chronic and post-COVID-19 cardiovascular complications. However, the magnitude of gene association was reduced for all arterial-related events (CAD, AF, and ISS), suggesting that distinctive pathogenic mechanisms may be involved when arterial disorders develop during COVID-19 infection^2,20^, such as the virus directly mediating heart injury by entering cardiomyocytes.^21,22^ Importantly, the polygenic risk for conventional VTE was remarkably retained for COVID-19-related VTE, which echo the finding in our previous study showing that the monogenic such as factor V Leiden mutation also consistently predisposed post-COVID-19 VTE complications.^6^

The role of genetics in variable COVID-19 representations is not yet well-understood. Although a large genome-wide association study (GWAS) was performed for COVID-19, it focused on SARS-COV-2-induced critical respiratory complications.^23,24^. We leveraged well-developed PRSs and proved that human polygenic variations affect CVE manifestations after COVID-19, beyond the severity of COVID-19 disease. There is also a lack of evidence for the potential beneficial effects of healthy lifestyles on reducing the cardiovascular disease burden in COVID-19. Although many studies have reported that lifestyle factors affected chronic cardiovascular conditions independently of individuals’ genetic background before the pandemic,^15,27^ little is known whether this remains the case in COVID-19-related cardiovascular complications. Some studies have observed lower risks of severe COVID-19 among people adopting more favourable behaviours.^25,26^ However, these studies concentrated on general health utilization outcomes such as hospital or ICU admissions, therefore limiting the specificity of the observed associations, particularly life-threatening CVE.

### Clinical and public health implications

Our findings have implications for clinical responses and public health preparedness against the ongoing COVID-19 pandemic. At the individual level, compared with known general risk factors such as demographic characteristics (e.g., age and sex) and clinical risk factors (e.g., obesity and hypertension), genetic factors might inform more tailored treatment choices to prevent specific COVID-19 complications. For instance, disease-specific PRSs could help doctors identify patients with high genetic risk for arterial thrombosis who would benefit from platelet inhibitors or identify those with high genetic risk for venous thrombosis such as VTE and prioritise them for coagulation cascade suppression therapy.^3,28^ Such specific PRSs cannot be achieved using traditional clinical factors alone, such as age, as they were associated with both high VTE risk and high adverse events related to pharmaceutical therapy such as antithrombotics.

Over the past 10 to 15 years, global interest, efforts, and controversies have surrounded PRSs’ clinical utility for the primary prevention of non-communicable cardiovascular diseases, such as predicting 10-year risk in the general population.^9,11,29^ The potential of a PRS could be magnified in patients with COVID-19 as they have a substantially increased CVE risk, particularly during the initial illness. If a PRS was calculated for everyone at birth and held as part of their health records,^30^ it could have been used as easily as demographic determinants like age and sex to refine existing approaches to defining subgroups who are particularly vulnerable to COVID-19, possibly providing more timely, personalised shielding advice. Even a small or modest improvement in stratification accuracy might lead to a sizeable population changing their COVID-19 vulnerability category.

Although the genetic risk for post-COVID-19 CVE is inherited, our study showed that acquired healthy behaviours could offset this risk. The US Preventive Services Task Force updated its guidelines in 2022, recommending behavioural counselling for cardiovascular disease prevention for all adults aged 18 years and older.^12,14^ Our data support this recommendation by showing that a composite healthy lifestyle can also benefit acute CVE outcomes after COVID-19, regardless of genetic risk. This benefit should be emphasized in future lifestyle intervention programmes during the ongoing pandemic.

### Strengths and limitations

Our study benefitted from a large population-based cohort, standardized genotyping, quality-controlled genetic data, powered and validated PRS estimates, well-defined measurements of a range of lifestyle factors, PCR-confirmed COVID-19 infection, and reliable and complete linkages to cardiovascular disease outcomes, which together enable these novel findings.

However, several study limitations should be considered. Our PRS was initially built to quantify polygenic risk for any adult-onset cardiovascular disease. It may not reflect the maximum possible genetic contribution to COVID-19 cardiovascular complications, especially given the likelihood of distinct pathological mechanisms involving virus-induced cardiovascular events. Future GWAS studies explicitly designed for COVID-19-related CVE could inform the development of a bespoke PRS and improve predictive performance.

Observational studies that use routinely collected data to ascertain disease outcomes may record overdiagnoses for COVID-19 patients. The ICD records of some clinical events, such as hypertension or diabetes, immediately after COVID-19 infection could be duplicate records of historical conditions instead of a new or activated disease status. However, all of the cardiovascular disease subtypes except for CAD used as outcomes in this study appeared to be temporary and potentially life-threatening. They are unlikely to be coded for without justification in actual clinical practice. Our sensitivity analyses using only incident or hospital-admission-specific CVE also produced findings consistent with the main analysis, precluding this concern.

Demonstrating statistical significance does not guarantee that the PRS is able to provide additional predictive information up on the existing clinical factors only based cardiovascular models, as previous studies have frequently found little agreement between statistical association and predictive performance.^31,32^ More modelling research is urgently needed to fill this evidence gap in the contexts of COVID-19.

We used lifestyle behaviour data collected 10 years ago as a surrogate for current lifestyle habits at the time of infection, which is likely subject to misclassification and may have biased any genuine associations toward the null. Reassuringly, all participants at the time of recruitment were middle-aged or older adults whose lifestyle habits should have been well established, suggesting that their habits are likely to have remained consistent over years between recruitment and infection.

Participants in UK Biobank represent a generally healthier population than the general population of the UK and are mostly of European ancestry, which may limit our findings’ generalisability beyond this population.

## Conclusions

Individuals’ genetic predisposition to cardiovascular disease, in the form of a PRS, was associated with a short-term risk of CAD, AF, and VTE complications after COVID-19, but not ISS. Additionally, the incidence of all studied arterial events was substantially lower among those adhering to a healthy lifestyle, independent of their genetic risk. These findings demonstrate that a person’s genetic background underpins how COVID-19 affects their cardiovascular system, but suggest that healthy lifestyle interventions can alleviate the elevated population cardiovascular disease burden during the ongoing pandemic.

## Online Methods

### Data sources and COVID-19 population

UK Biobank is a large-scale, population-based prospective cohort of over 500,000 individuals aged 40 to 69 years at recruitment between 2006 and 2010 from across the United Kingdom.^33^ Extensive information was collected through questionnaires and physical measurements at 22 assessment centres. Affymetrix performed genotype calling based on two closely related purpose-designed arrays.^34^ Follow-up health outcomes were identified through linkage to various electronic health records, covering national primary and secondary care, disease and mortality registries.^35^ The reliability, accuracy and completeness of capturing disease conditions using this approach, especially cardiovascular diseases, have been validated in previous studies.^36,37^ To enable urgent research into COVID-19, additional data from Public Health England’s Second Generation Surveillance System has recently been linked to all UK Biobank participants using a bespoke algorithm.^38^ This information includes whether their SARS-CoV-2 infection status has been confirmed with polymerase chain reaction (PCR) testing.

In this study, we enrolled a cohort of participants from UK Biobank with COVID-19 from England who had a positive PCR testing result between March 1, 2020, and September 30, 2021. For one sensitivity analysis, we used all participants in the full UK Biobank population who were alive on March 1, 2020. Participants with missing age, sex, BMI, socioeconomic status, lifestyle factors, and genotyping information at baseline were excluded.

### Polygenic risk scores

In May 2022, UK Biobank released the first version of polygenic risk scores for 28 diseases, proving their predictive ability to outperform currently published PRS (see Supplementary Methods).^39^ Two sets of PRS, standard and enhanced, were developed and validated. The standard score was developed based only on non-UK-Biobank populations and then calculated for all individuals in UK Biobank. The development of the enhanced score used some UK Biobank participants, with the score then computed for those remaining to avoid the risk of overfitting.^40^ In this study, we used the standard PRS for coronary artery disease (CAD), atrial fibrillation (AF), ischaemic stroke (ISS), and venous thromboembolic disease (VTE) in the primary analysis and the enhanced PRS in one of the sensitivity analyses. The continuous PRS used in this study was also categorized as high-risk (fifth quintile), intermediate-risk (2-4 quintiles), and low-risk (lowest quintile) to enable comparisons with previous studies.^27,41^

### Definition of healthy lifestyles

We defined a composite healthy lifestyle index by combining data for nine lifestyle components based on a published study^42^: smoking status, alcohol drinking, physical activity, television viewing time, sleep duration, intake of fruit and vegetable, intake of oily fish, intake of red meat, and intake of processed meat. Each lifestyle factor was assigned 0 points if healthy and 1 point if unhealthy. For example, participants with “alcohol drinking daily or almost daily” scored 1 point for the alcohol drinking element. We then summed all lifestyle factors, with a score of 0-4 classified as combined healthy lifestyle and a score of 5-9 classified as unhealthy lifestyle. More details on defining each beneficial lifestyle factor are provided in **Supplemental Methods**.

### Post-COVID-19 CVE

Among participants with COVID-19, we defined the first infection as the index date and followed up for 90 days. We studied four major CVEs (AF, CAD, ISS, and VTE) that were frequently reported as COVID-19-related cardiovascular complications, through linkage to hospital admissions data. International Classification of Diseases 10^th^ Revision (ICD-10) codes were used to capture clinical outcomes and are presented in **Supplemental Methods**. These ICD-10 codes were the same as those initially used for PRS development to minimize the impact of variation in disease phenotypes between our and previous studies.^40^ Data in this study were censored on September 30, 2021.

### Statistical Analyses

We used the Cox proportional hazard (PH) model to assess the associations between each PRS and its corresponding CVE outcome. PH assumptions were checked based on Schoenfeld residuals and were satisfied. The hazard ratio (HR) and 95% confidence interval (CI) for a continuous PRS (per 1 standard deviation [SD] increase) were derived by adjusting for age, sex, education level (mapped to the international standard for classification of education, see **Supplemental Methods**), index of multiple deprivations (IMD, a continuous summary deprivation measurement used in England that contains crime, education, employment, health, housing, income, and living environment),^43^ genotyping batch, and the first ten principal components of genetic ancestry. To avoid overadjustment, we did not adjust for previous CVE conditions (which, for example, can mediate the genetic effects on post-COVID-19 CVE outcomes).

We calculated the PRS-CVE association and stratified it by clinically relevant features, including age (≥ 65 years or < 65 years), sex, body mass index (≥ 30 or < 30), ethnicity (White or other ethnic groups), recent antithrombotic medication (yes or no), setting for a positive PRC test (inpatient or outpatient), and SARS-COV-2 infection type (breakthrough infection after two-dose vaccination or non-breakthrough infection). Multiplicative interactions between PRS and the stratification variables were tested, and *P*-values are reported. Restricted cubic splines were used to examine possible nonlinear associations for the continuous PRS.^44^ Categorical genetic risk was analyzed separately and depicted using the Kaplan–Meier method.

We used the same multivariable Cox regression model for the lifestyle factor in the overall cohort and across the PRS categories. We calculated the prevented fraction for the population (PFP) according to the formula: PFP = Pe (1–HR), where Pe was the prevalence of the unhealthy lifestyle.^45^ We estimated the combined effect of genetics and lifestyle (six categories with low genetic risk and healthy lifestyle as reference group) on those CVE outcomes that were statistically associated with both genetic and lifestyle factors.

We performed several sensitivity analyses for genetic risk. First, we examined the associations among the sub-cohort of participants with an enhanced PRS recorded in UK Biobank. Second, we studied incident CVE by excluding participants with a historical study outcome before the index date. Third, we defined the CVE outcomes based on the primary diagnosis alone, which reflects the main cause of hospital admission. Fourth, we performed a negative control outcome experiment using the association between PRS and type 2 diabetes among the COVID-19 cohort and a positive control outcome experiment on the association between PRS and CVE among UK Biobank participants without evidence of COVID-19. The positive and negative control experiments were designed to detect residual confounding and any spurious bias related to study design, cohort building, analytic approach, or outcome ascertainment.

The analyses were performed using R software version 4.1.2. All statistical tests were 2-sided. A 95% CI that did not contain unity was considered statistically significant.

## Data Availability

All data produced in the present study are available upon reasonable request to the authors

## Supplementary Results

**sTable 1:** Characteristics of the whole UK Biobank population and of participants with COVID-19, stratified by genetic risk of coronary artery disease.

| Participant characteristics | All eligible UK Biobank participants<sup>2</sup> | Participants with COVID-19 |  |  |  |
| --- | --- | --- | --- | --- | --- |
|  |  | All | Low genetic risk | Intermediate genetic risk | High genetic risk |
| Number | 408395 | 25335 | 5067 | 15201 | 5067 |
| Mean age, year (SD) | 68.10 (8.10) | 65.99 (8.53) | 66.08 (8.49) | 66.05 (8.55) | 65.74 (8.54) |
| sex, No. (%) |  |  |  |  |  |
| Female | 225451 (55.2) | 13339 (52.7) | 2656 (52.4) | 8044 (52.9) | 2639 (52.1) |
| Male | 182944 (44.8) | 11996 (47.3) | 2411 (47.6) | 7157 (47.1) | 2428 (47.9) |
| Ethnicity, No. (%) |  |  |  |  |  |
| White | 382169 (93.6) | 3893 (15.4) | 820 (16.2) | 2244 (14.8) | 829 (16.4) |
| Other ethnic groups | 26226 ( 6.4) | 21442 (84.6) | 4247 (83.8) | 12957 (85.2) | 4238 (83.6) |
| Obesity, No. (%) |  |  |  |  |  |
| BMI < 30 | 311205 (76.2) | 17984 (71.0) | 3695 (72.9) | 10734 (70.6) | 3555 (70.2) |
| BMI ≥ 30 | 97190 (23.8) | 7351 (29.0) | 1372 (27.1) | 4467 (29.4) | 1512 (29.8) |
| Education categories<sup>3</sup>, No. (%) |  |  |  |  |  |
| I | 73987 (18.1) | 4863 (19.2) | 880 (17.4) | 2932 (19.3) | 1051 (20.7) |
| II | 110969 (27.2) | 7999 (31.6) | 1526 (30.1) | 4839 (31.8) | 1634 (32.2) |
| III | 45409 (11.1) | 2613 (10.3) | 553 (10.9) | 1546 (10.2) | 514 (10.1) |
| IV | 20765 ( 5.1) | 1194 ( 4.7) | 260 ( 5.1) | 705 ( 4.6) | 229 ( 4.5) |
| V | 157265 (38.5) | 8666 (34.2) | 1848 (36.5) | 5179 (34.1) | 1639 (32.3) |
| Index of multiple deprivation, mean (SD) | 17.51 (13.87) | 20.62 (14.85) | 20.61 (14.97) | 20.47 (14.69) | 21.07 (15.18) |
| **Individual lifestyle factors, No. (%)** |  |  |  |  |  |
| Smoking status |  |  |  |  |  |
| Never or past | 368471 (90.2) | 22590 (89.2) | 4543 (89.7) | 13565 (89.2) | 4482 (88.5) |
| Current | 39924 ( 9.8) | 2745 (10.8) | 524 (10.3) | 1636 (10.8) | 585 (11.5) |
| Alcohol intake |  |  |  |  |  |
| ≤ 4 times week | 325341 (79.7) | 21062 (83.1) | 4143 (81.8) | 12671 (83.4) | 4248 (83.8) |
| Daily or almost daily | 83054 (20.3) | 4273 (16.9) | 924 (18.2) | 2530 (16.6) | 819 (16.2) |
| Physical activity |  |  |  |  |  |
| ≥150 min per week of MIPA or ≥75 min per week of VIPA | 345208 (84.5) | 21194 (83.7) | 4243 (83.7) | 12751 (83.9) | 4200 (82.9) |
| <150 min per week of MIPA & <75 min per week of VIPA | 63187 (15.5) | 4141 (16.3) | 824 (16.3) | 2450 (16.1) | 867 (17.1) |
| Television viewing time |  |  |  |  |  |
| < 4 h/day | 293476 (71.9) | 17880 (70.6) | 3661 (72.3) | 10747 (70.7) | 3472 (68.5) |
| ≥ 4 h/day | 114919 (28.1) | 7455 (29.4) | 1406 (27.7) | 4454 (29.3) | 1595 (31.5) |
| Sleep duration |  |  |  |  |  |
| ≥7 & ≤9h/day | 301723 (73.9) | 18121 (71.5) | 3667 (72.4) | 10888 (71.6) | 3566 (70.4) |
| <7 or >9h/day | 106672 (26.1) | 7214 (28.5) | 1400 (27.6) | 4313 (28.4) | 1501 (29.6) |
| Fruit and vegetable intake |  |  |  |  |  |
| ≥ 400 g/day | 327452 (80.2) | 19456 (76.8) | 3879 (76.6) | 11677 (76.8) | 3900 (77.0) |
| <400 g/ day | 80943 (19.8) | 5879 (23.2) | 1188 (23.4) | 3524 (23.2) | 1167 (23.0) |
| Oily fish intake |  |  |  |  |  |
| ≥1 portion/week | 230406 (56.4) | 13047 (51.5) | 2638 (52.1) | 7775 (51.1) | 2634 (52.0) |
| <1 portion/week | 177989 (43.6) | 12288 (48.5) | 2429 (47.9) | 7426 (48.9) | 2433 (48.0) |
| Red meat intake |  |  |  |  |  |
| ≤3 portion/week | 188229 (46.1) | 13118 (51.8) | 2545 (50.2) | 7883 (51.9) | 2690 (53.1) |
| >3 portion/week | 220166 (53.9) | 12217 (48.2) | 2522 (49.8) | 7318 (48.1) | 2377 (46.9) |
| Processed meat intake |  |  |  |  |  |
| ≤1 portion/week | 282323 (69.1) | 16716 (66.0) | 3354 (66.2) | 9971 (65.6) | 3391 (66.9) |
| >1 portion/week | 126072 (30.9) | 8619 (34.0) | 1713 (33.8) | 5230 (34.4) | 1676 (33.1) |
| Combined lifestyle, No. (%) |  |  |  |  |  |
| Healthy (0-4 unhealthy factors) | 377279 (92.4) | 23151 (91.4) | 4627 (91.3) | 13889 (91.4) | 4635 (91.5) |
| Unhealthy (5-9 unhealthy factors) | 31116 ( 7.6) | 2184 ( 8.6) | 440 ( 8.7) | 1312 ( 8.6) | 432 ( 8.5) |

**sTable 2:** Characteristics of the whole UK Biobank population and of participants with COVID-19, stratified by genetic risk of ischaemic stroke.

| Participant characteristics | All eligible UK Biobank participants<sup>2</sup> | Participants with COVID-19 |  |  |  |
| --- | --- | --- | --- | --- | --- |
|  |  | All | Low genetic risk | Intermediate genetic risk | High genetic risk |
| Number | 408395 | 25335 | 5067 | 15201 | 5067 |
| Mean age, year (SD) | 68.10 (8.10) | 65.99 (8.53) | 65.95 (8.53) | 66.06 (8.54) | 65.81 (8.53) |
| sex, No. (%) |  |  |  |  |  |
| Female | 225451 (55.2) | 13339 (52.7) | 2665 (52.6) | 8012 (52.7) | 2662 (52.5) |
| Male | 182944 (44.8) | 11996 (47.3) | 2402 (47.4) | 7189 (47.3) | 2405 (47.5) |
| Ethnicity, No. (%) |  |  |  |  |  |
| White | 382169 (93.6) | 3893 (15.4) | 886 (17.5) | 2250 (14.8) | 757 (14.9) |
| Other ethnic groups | 26226 ( 6.4) | 21442 (84.6) | 4181 (82.5) | 12951 (85.2) | 4310 (85.1) |
| Obesity, No. (%) |  |  |  |  |  |
| BMI < 30 | 311205 (76.2) | 17984 (71.0) | 3789 (74.8) | 10835 (71.3) | 3360 (66.3) |
| BMI ≥ 30 | 97190 (23.8) | 7351 (29.0) | 1278 (25.2) | 4366 (28.7) | 1707 (33.7) |
| Education categories<sup>3</sup>, No. (%) |  |  |  |  |  |
| I | 73987 (18.1) | 4863 (19.2) | 866 (17.1) | 2909 (19.1) | 1088 (21.5) |
| II | 110969 (27.2) | 7999 (31.6) | 1518 (30.0) | 4867 (32.0) | 1614 (31.9) |
| III | 45409 (11.1) | 2613 (10.3) | 568 (11.2) | 1531 (10.1) | 514 (10.1) |
| IV | 20765 ( 5.1) | 1194 ( 4.7) | 241 ( 4.8) | 705 ( 4.6) | 248 ( 4.9) |
| V | 157265 (38.5) | 8666 (34.2) | 1874 (37.0) | 5189 (34.1) | 1603 (31.6) |
| Index of multiple deprivation, mean (SD) | 17.51 (13.87) | 20.62 (14.85) | 20.41 (14.80) | 20.45 (14.73) | 21.32 (15.23) |
| **Individual lifestyle factors, No. (%)** |  |  |  |  |  |
| Smoking status |  |  |  |  |  |
| Never or past | 368471 (90.2) | 22590 (89.2) | 4571 (90.2) | 13548 (89.1) | 4471 (88.2) |
| Current | 39924 ( 9.8) | 2745 (10.8) | 496 ( 9.8) | 1653 (10.9) | 596 (11.8) |
| Alcohol intake |  |  |  |  |  |
| ≤ 4 times week | 325341 (79.7) | 21062 (83.1) | 4188 (82.7) | 12618 (83.0) | 4256 (84.0) |
| Daily or almost daily | 83054 (20.3) | 4273 (16.9) | 879 (17.3) | 2583 (17.0) | 811 (16.0) |
| Physical activity |  |  |  |  |  |
| ≥150 min per week of MIPA or ≥75 min per week of VIPA | 345208 (84.5) | 21194 (83.7) | 4284 (84.5) | 12766 (84.0) | 4144 (81.8) |
| <150 min per week of MIPA & <75 min per week of VIPA | 63187 (15.5) | 4141 (16.3) | 783 (15.5) | 2435 (16.0) | 923 (18.2) |
| Television viewing time |  |  |  |  |  |
| < 4 h/day | 293476 (71.9) | 17880 (70.6) | 3721 (73.4) | 10733 (70.6) | 3426 (67.6) |
| ≥ 4 h/day | 114919 (28.1) | 7455 (29.4) | 1346 (26.6) | 4468 (29.4) | 1641 (32.4) |
| Sleep duration |  |  |  |  |  |
| ≥7 & ≤9h/day | 301723 (73.9) | 18121 (71.5) | 3687 (72.8) | 10912 (71.8) | 3522 (69.5) |
| <7 or >9h/day | 106672 (26.1) | 7214 (28.5) | 1380 (27.2) | 4289 (28.2) | 1545 (30.5) |
| Fruit and vegetable intake |  |  |  |  |  |
| ≥ 400 g/day | 327452 (80.2) | 19456 (76.8) | 3904 (77.0) | 11704 (77.0) | 3848 (75.9) |
| <400 g/ day | 80943 (19.8) | 5879 (23.2) | 1163 (23.0) | 3497 (23.0) | 1219 (24.1) |
| Oily fish intake |  |  |  |  |  |
| ≥1 portion/week | 230406 (56.4) | 13047 (51.5) | 2594 (51.2) | 7880 (51.8) | 2573 (50.8) |
| <1 portion/week | 177989 (43.6) | 12288 (48.5) | 2473 (48.8) | 7321 (48.2) | 2494 (49.2) |
| Red meat intake |  |  |  |  |  |
| ≤3 portion/week | 188229 (46.1) | 13118 (51.8) | 2473 (48.8) | 7915 (52.1) | 2730 (53.9) |
| >3 portion/week | 220166 (53.9) | 12217 (48.2) | 2594 (51.2) | 7286 (47.9) | 2337 (46.1) |
| Processed meat intake |  |  |  |  |  |
| ≤1 portion/week | 282323 (69.1) | 16716 (66.0) | 3337 (65.9) | 10059 (66.2) | 3320 (65.5) |
| >1 portion/week | 126072 (30.9) | 8619 (34.0) | 1730 (34.1) | 5142 (33.8) | 1747 (34.5) |
| Combined lifestyle, No. (%) |  |  |  |  |  |
| Healthy (0-4 unhealthy factors) | 377279 (92.4) | 23151 (91.4) | 4641 (91.6) | 13924 (91.6) | 4586 (90.5) |
| Unhealthy (5-9 unhealthy factors) | 31116 ( 7.6) | 2184 ( 8.6) | 426 ( 8.4) | 1277 ( 8.4) | 481 ( 9.5) |

**sTable 3:** Characteristics of the whole UK Biobank population and of participants with COVID-19, stratified by genetic risk of venous thromboembolism.

| Participant characteristics | All eligible UK Biobank participants<sup>2</sup> | Participants with COVID-19 |  |  |  |
| --- | --- | --- | --- | --- | --- |
|  |  | All | Low genetic risk | Intermediate genetic risk | High genetic risk |
| Number | 408395 | 25335 | 5067 | 15201 | 5067 |
| Mean age, year (SD) | 68.10 (8.10) | 65.99 (8.53) | 65.87 (8.57) | 66.05 (8.54) | 65.95 (8.47) |
| sex, No. (%) |  |  |  |  |  |
| Female | 225451 (55.2) | 13339 (52.7) | 2660 (52.5) | 7971 (52.4) | 2708 (53.4) |
| Male | 182944 (44.8) | 11996 (47.3) | 2407 (47.5) | 7230 (47.6) | 2359 (46.6) |
| Ethnicity, No. (%) |  |  |  |  |  |
| White | 382169 (93.6) | 3893 (15.4) | 789 (15.6) | 2296 (15.1) | 808 (15.9) |
| Other ethnic groups | 26226 ( 6.4) | 21442 (84.6) | 4278 (84.4) | 12905 (84.9) | 4259 (84.1) |
| Obesity, No. (%) |  |  |  |  |  |
| BMI < 30 | 311205 (76.2) | 17984 (71.0) | 3630 (71.6) | 10805 (71.1) | 3549 (70.0) |
| BMI ≥ 30 | 97190 (23.8) | 7351 (29.0) | 1437 (28.4) | 4396 (28.9) | 1518 (30.0) |
| Education categories<sup>3</sup>, No. (%) |  |  |  |  |  |
| I | 73987 (18.1) | 4863 (19.2) | 904 (17.8) | 2949 (19.4) | 1010 (19.9) |
| II | 110969 (27.2) | 7999 (31.6) | 1632 (32.2) | 4750 (31.2) | 1617 (31.9) |
| III | 45409 (11.1) | 2613 (10.3) | 536 (10.6) | 1546 (10.2) | 531 (10.5) |
| IV | 20765 ( 5.1) | 1194 ( 4.7) | 252 ( 5.0) | 715 ( 4.7) | 227 ( 4.5) |
| V | 157265 (38.5) | 8666 (34.2) | 1743 (34.4) | 5241 (34.5) | 1682 (33.2) |
| Index of multiple deprivation, mean (SD) | 17.51 (13.87) | 20.62 (14.85) | 20.31 (14.58) | 20.68 (14.96) | 20.72 (14.78) |
| **Individual lifestyle factors, No. (%)** |  |  |  |  |  |
| Smoking status |  |  |  |  |  |
| Never or past | 368471 (90.2) | 22590 (89.2) | 4540 (89.6) | 13510 (88.9) | 4540 (89.6) |
| Current | 39924 ( 9.8) | 2745 (10.8) | 527 (10.4) | 1691 (11.1) | 527 (10.4) |
| Alcohol intake |  |  |  |  |  |
| ≤ 4 times week | 325341 (79.7) | 21062 (83.1) | 4224 (83.4) | 12591 (82.8) | 4247 (83.8) |
| Daily or almost daily | 83054 (20.3) | 4273 (16.9) | 843 (16.6) | 2610 (17.2) | 820 (16.2) |
| Physical activity |  |  |  |  |  |
| ≥150 min per week of MIPA or ≥75 min per week of VIPA | 345208 (84.5) | 21194 (83.7) | 4253 (83.9) | 12750 (83.9) | 4191 (82.7) |
| <150 min per week of MIPA & <75 min per week of VIPA | 63187 (15.5) | 4141 (16.3) | 814 (16.1) | 2451 (16.1) | 876 (17.3) |
| Television viewing time |  |  |  |  |  |
| < 4 h/day | 293476 (71.9) | 17880 (70.6) | 3609 (71.2) | 10697 (70.4) | 3574 (70.5) |
| ≥ 4 h/day | 114919 (28.1) | 7455 (29.4) | 1458 (28.8) | 4504 (29.6) | 1493 (29.5) |
| Sleep duration |  |  |  |  |  |
| ≥7 & ≤9h/day | 301723 (73.9) | 18121 (71.5) | 3602 (71.1) | 10930 (71.9) | 3589 (70.8) |
| <7 or >9h/day | 106672 (26.1) | 7214 (28.5) | 1465 (28.9) | 4271 (28.1) | 1478 (29.2) |
| Fruit and vegetable intake |  |  |  |  |  |
| ≥ 400 g/day | 327452 (80.2) | 19456 (76.8) | 3880 (76.6) | 11663 (76.7) | 3913 (77.2) |
| <400 g/ day | 80943 (19.8) | 5879 (23.2) | 1187 (23.4) | 3538 (23.3) | 1154 (22.8) |
| Oily fish intake |  |  |  |  |  |
| ≥1 portion/week | 230406 (56.4) | 13047 (51.5) | 2582 (51.0) | 7893 (51.9) | 2572 (50.8) |
| <1 portion/week | 177989 (43.6) | 12288 (48.5) | 2485 (49.0) | 7308 (48.1) | 2495 (49.2) |
| Red meat intake |  |  |  |  |  |
| ≤3 portion/week | 188229 (46.1) | 13118 (51.8) | 2623 (51.8) | 7879 (51.8) | 2616 (51.6) |
| >3 portion/week | 220166 (53.9) | 12217 (48.2) | 2444 (48.2) | 7322 (48.2) | 2451 (48.4) |
| Processed meat intake |  |  |  |  |  |
| ≤1 portion/week | 282323 (69.1) | 16716 (66.0) | 3338 (65.9) | 10038 (66.0) | 3340 (65.9) |
| >1 portion/week | 126072 (30.9) | 8619 (34.0) | 1729 (34.1) | 5163 (34.0) | 1727 (34.1) |
| Combined lifestyle, No. (%) |  |  |  |  |  |
| Healthy (0-4 unhealthy factors) | 377279 (92.4) | 23151 (91.4) | 4646 (91.7) | 13880 (91.3) | 4625 (91.3) |
| Unhealthy (5-9 unhealthy factors) | 31116 ( 7.6) | 2184 ( 8.6) | 421 ( 8.3) | 1321 ( 8.7) | 442 ( 8.7) |

**SFigure 1:**
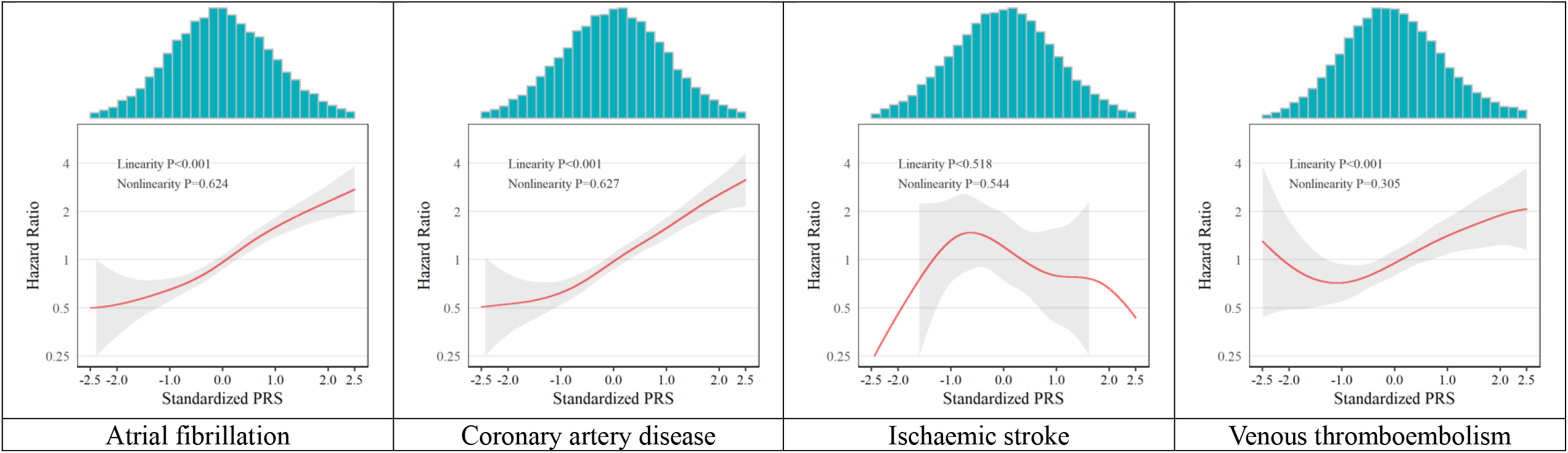
Dose-response associations between continuous polygenic risk scores (PRSs) and study outcomes. If the confidence interval of the hazard ratio was larger than 4 or smaller than 0.25, it was removed.

**sTable 4:** Sensitivity analyses.

|  | **Atrial fibrillation** | **Coronary artery disease** | **Ischaemic stroke** | **Venous thromboembolism** |
| --- | --- | --- | --- | --- |
| **Sensitivity results** |  |  |  |  |
| Use of enhanced PRS | 1.67 (1.35 to 2.06) | 1.51 (1.17 to 1.95) | 0.81 (0.43 to 1.50) | 1.32 (0.98 to 1.78) |
| Incident cardiovascular and thromboembolic events | 1.33 (1.11 to 1.59) | 1.19 (0.87 to 1.63) | 0.84 (0.55 to 1.27) | 1.32 (1.11 to 1.55) |
| Hospital admission diagnosis | 1.31 (0.89 to 1.92) | 1.21 (0.71 to 2.08) | 0.87 (0.56 to 1.36) | 1.52 (1.14 to 2.02) |
| COVID-19-unrealtd CVE (Positive control outcome) | 1.65 (1.58 to 1.72) | 1.56 (1.47 to 1.65) | 1.32 (1.13 to 1.53) | 1.41 (1.24 to 1.61) |
| Diabetes (Negative control outcome) | 0.82 (0.69 to 0.97) | 0.98 (0.83 to 1.15) | 1.13 (0.96 to 1.34) | 1.09 (0.93 to 1.29) |

## Supplementary Methods

**Predictive performance of the UK Biobank PRS Release (the black point) against published comparator PRSs (the colourful point)**.

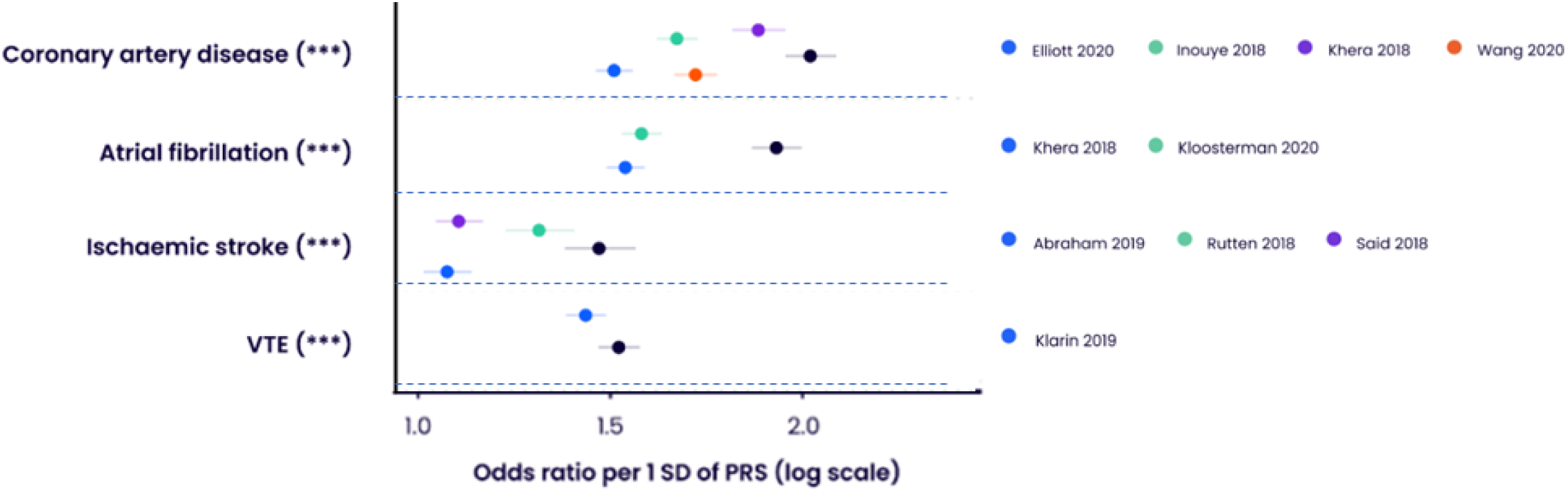
(Citation: Deborah J. Thompson, Preprint medRxiv DOI: https://doi.org/10.1101/2022.06.16.22276246, 2022)

**UK Biobank self-reported highest qualification mapped to the International Standard Classification of Education (ISCED)**.

| **UK Biobank self-reported highest qualification** | **Years of education** | **ISCED** |
| --- | --- | --- |
| Prefer not to answer | 7 | 1 |
| None of the above | 7 | 1 |
| CSEs or equivalent | 10 | 2 |
| O levels/GCSEs or equivalent | 10 | 2 |
| A levels/AS levels or equivalent | 13 | 3 |
| Other professional qualifications eg: nursing, teaching | 15 | 4 |
| NVQ or HND or HNC or equivalent | 19 | 5 |
| College or University degree | 20 | 5 |

**Variables in UK Biobank used to create the lifestyle score in this study.**

| **Lifestyle factor** | **Questionnaire** | **Healthy (0)** | **Unhealthy (1)** |
| --- | --- | --- | --- |
| **1. Smoking status** | "Do you smoke tobacco now?" and "In the past, how often have you smoked tobacco?" | past or never smoker | current |
| **2. Alcohol intake** | "About how often do you drink alcohol?" | ≤ 4 times week | Daily or almost daily |
| **3. Physical activity** | IPAQ short form<sup>2</sup>– total time walking or moderate and vigorous-intensity PA in previous week * | ≥150 min/week moderate or ≥ 75 min/week vigorous PA | < 75 min/week vigorous PA |
| **4. TV viewing /sedentary time** | "In a typical day, how many hours do you spend watching TV?" † | < 4 h/day | ≥ 4 h/day |
| **5. Sleep time** | "About how many hours sleep do you get in every 24 hours?" | >7 or <9h/day | <7 or >9h/day |
| **Dietary characteristics** | Individual dietary components contributed directly to lifestyle score.Φ |  |  |
| **6. Fruit and vegetable intake** | "About how many of .... would you eat per day?" Separate questions for pieces of fresh and dried fruit, tablespoons of salad or cooked/raw vegetables. Combined and converted to g/day (1 portion = 80 g) | ≥ 400 g/day | <400 g/ day |
| **7. Oily fish intake** | "How often do you eat oily fish? (e.g. sardines, salmon, mackerel, herring)" | ≥1 portion/week | <1 portion/week |
| **8. Red meat intake** | "How often do you eat...?" Separate questions for Beef / lamb or mutton / pork (excluding processed meats such as ham or bacon). Red meat included due to clear link between red meat and mortality.<sup>3</sup> | ≤3 portion/week | >3 portion/week |
| **9. Processed meat intake** | "How often do you eat processed meats (such as bacon, ham, sausages, meat pies, kebabs, burgers, chicken nuggets)?" | ≤1 portion/week | >1 portion/week |

Participants were classified as “not at risk/healthy” or “at risk/unhealthy” based on their responses to questions for each lifestyle factor. \*UK Biobank physical activity (PA) data were analyzed in accordance with the International Physical Activity Questionnaire (IPAQ) scoring protocol, with total physical activity computed as the sum of walking, moderate, and vigorous activity, measured as metabolic equivalents (MET-hours/week). † UK Biobank participants were asked about work-related sitting time and were not asked about discretionary/leisure time sedentary behavior. Therefore, TV viewing time, which is strongly associated with mortality, was used instead of sedentary time. UK Biobank participants were asked separately about time spent using a computer, excluding computer time at work. This information was used to check for implausible values for TV viewing time. Φ National dietary guidelines were used as the basis for the dietary components.

*(Developed by Hamish M, Lancet Public Health 2018)*

**ICD-10 codes originally used to develop the polygenic risk score and define cardiovascular and thromboembolic events in this study.**

| **Outcomes** | **ICD-10 codes** |
| --- | --- |
| Atrial fibrillation | I48 |
| Coronary artery disease | I21, I22, I23, I24.1, I25.2 |
| Ischaemic stroke | I63, I64 |
| Venous thromboembolic disease | I81, I82, I26, O22.3, O87.1, O08.2 |

## Declaration of interests

DPA’s department has received grant/s from Amgen, Chiesi-Taylor, Lilly, Janssen, Novartis, and UCB Biopharma. His research group has received consultancy fees from Astra Zeneca and UCB Biopharma. Amgen, Astellas, Janssen, Synapse Management Partners and UCB Biopharma have funded or supported training programmes organised by DPA’s department. Roger Paredes has participated in advisory boards for Gilead, MSD, ViiV Healthcare, Theratechnologies and Lilly. His institution has received research support from Gilead, MSD, and ViiV Healthcare.

### Ethical Approval

All participants provided written informed consent at the UKBB cohort recruitment. This study received ethical approval from the UKBB Ethics Advisory Committee (EAC) under application 65397.

### Data Sharing

Bonafide researchers can apply to use the UK Biobank dataset by registering and applying at http://ukbiobank.ac.uk/register-apply/. Any additional summary data generated and/or analysed during the current study are available from the corresponding author upon reasonable request.

